# Contact Tracing Evaluation for COVID-19 Transmission during the Reopening Phase in a Rural College Town

**DOI:** 10.1101/2020.06.24.20139204

**Authors:** Sifat Afroj Moon, Caterina Scoglio

## Abstract

Contact tracing can play a vital role in controlling human-to-human transmission of a highly contagious disease such as COVID-19. To investigate the benefits and costs of contact tracing, we develop an individual-based contact-network model and a susceptible-exposed-infected-confirmed (SEIC) epidemic model for the stochastic simulations of COVID-19 transmission. We estimate the unknown parameters (reproductive ratio *R*_0_ and confirmed rate *δ*_2_) by using observed confirmed case data. After a two month-lockdown, states in the USA have started the reopening process. We investigate for four different reopening situations: under “stay-at-home” order or no reopening, 25 % reopening, 50 % reopening, and 75 % reopening. We model contact tracing in a two-layer network by modifying the basic SEIC epidemic model. The two-layer network is composed by the contact network in the first layer and the tracing network in the second layer. Since the full contact list of an infected individual patient can be hard to obtain, then we consider different fractions of contacts from 60% to 5%. The goal of this paper is to assess the effectiveness of contact tracing to control the COVID-19 spreading during the initial phase of the reopening process of a rural college town.

In this research, we assess the benefits and cost of contact tracing as a key mitigation strategy to control the spreading of COVID-19. In terms of benefits, our simulation results show that increasing the fraction of traced contacts decreases the size of the epidemic. For example, tracing 20% of the contacts is enough for all four reopening scenarios to reduce the epidemic size by half. Considering the act of quarantining susceptible households as the contact tracing cost, we have observed an interesting phenomenon. When we increase the fraction of traced contacts from 5% to 20%, the number of quarantined susceptible people increases because each individual confirmed case is mentioning more contacts. However, when we increase the fraction of traced contacts from 20% to 60%, the number of quarantined susceptible people decreases because the increment of the mentioned contacts is balanced by a reduced number of confirmed cases. The outcomes of this research are valuable in the reopening process of the USA. Furthermore, the framework is generic enough to use any locations and for other diseases as well.

## I. INTRODUCTION

COVID-19 has affected the lives of billions of people in 2019-2020. The COVID-19 disease is caused by severe acute respiratory syndrome coronavirus 2 (SARS-CoV-2) and has caused a global health emergency. The world health organization (WHO) declared it as a Public Health Emergency of International Concern on January 30, 2020^1^. The number of confirmed reported cases by SARS-CoV-2 has been rising. On May 31, 2020, worldwide there were 5, 939, 234 laboratory-confirmed cases with 367, 255 deaths^2^.

Many countries issued a pandemic lockdown to slow down the spreading of COVID-19. In the United States, a ‘stay-at-home’ order was issued in many states. However, those pandemic lockdowns have a massive impact on the economy. All the States of the USA started reopening gradually from early May. Understanding the impact of mitigation strategies on the spreading dynamic of COVID-19 during the reopening phase of the USA is essential. In this work, we assess the impact of contact tracing under four reopening scenarios: 25% reopening, 50% reopening, 75% reopening, and 100% reopening.

Individual-based contact-network models are a powerful tool to model COVID-19 spreading due to its person-to-person spreading nature. In this work, we develop an individual-based network model for a college town, Manhattan, KS, where households represent nodes of the network. We select Manhattan, KS, as our study area, since it is a typical college town in a rural region of Kansas, the home of Kansas State University. There are 20, 439 occupied households in Manhattan, KS, according to census 2018^3^. The connections between two individual households represent the contact probabilities between the members of the households. The individual-based approach provides the flexibility to observe the local dynamic at the individual level. It also allows us to include in the model a mitigation strategy at the individual level, such as contact tracing.

To design an epidemic model for COVID-19 is challenging, as many epidemic features of the disease are yet to be investigated, such as, for example, the transmission rate, the pre-symptomatic transmission rate, and the percentage of the asymptomatic population. These uncertain characteristics make epidemic modeling challenging as the outcomes of the model are sensitive to the assumption made on the uncertainties. Therefore, we use a simple epidemic model with four compartments –susceptible-exposed-infected-confirmed (SEIC)– capable of imitating the COVID-19 transmission and flexible enough to cope with new information. This model has only two unknown parameters: the reproductive ratio *R*_0_, and the confirmed case rate or reporting rate *δ*_2_. We use confirmed COVID-19 cases from March 25, 2020 to May 4, 2020 in Manhattan, KS as data, and estimate the unknown parameters from data. We consider that a confirmed COVID-19 patient cannot spread the disease anymore except in his/her own household. In the spreading of COVID-19, there are pre-symptomatic and asymptomatic cases that do not show any sign of illness^4^. Besides, there is a strong possibility that infected cases not detected exist. In our epidemic model, those unreported cases are included indirectly through infected to confirmed transitions.

Since a vaccine is not available for COVID-19, contact tracing is a key mitigation strategy to control the spreading of COVID-19. Contact tracing is a mitigation strategy that aims at identifying people who may have come into contact with a patient. This mitigation strategy prevents further spreading by isolation of exposed people. The public health personnel have used contact tracing as a tool to control disease-spreading for a long time^5^. We implement the contact tracing strategy according to CDC guidance^6^ through a two-layer network model with a modified SEIC epidemic model. Contact tracing is effective at the early stage of an epidemic when there is a limited number of cases. We choose a college town, Manhattan (KS), for our study. Most college towns have a limited number of cases because educational institutes have been closed since early March 2020. Feasibility of contact tracing to control COVID-19 spreading was analyzed using a branching process stochastic simulation for three reproductive ratios *R*_0_ = 1.5, 2.5, and 3.5^7^. The authors find that sufficient contact tracing with quarantine can control a new outbreak of COVID-19. They mostly focus on the question of how much contacts need to be traced to control an epidemic for the three levels of reproductive ratio. However, this article neither explored the effectiveness of contact tracing for a specific location, nor investigated the cost of contact tracing.

In this research, we develop an individual-based network framework to assess the impact of contact-tracing in the reopening process in a college town of Kansas. To analyze the cost of contact-tracing represented by the number of quarantined susceptible people, we develop a contact network and estimate the reproductive ratio *R*_0_ and confirmed rate (infected to laboratory-confirmed transition) from observed confirmed case data in Manhattan KS. We use our individual-based network model and the estimated parameters to run simulations of COVID-19 transmission. We use our framework to understand the spreading of COVID-19 and assess the contact-tracing strategy in the different reopening situations and different levels of tracing contacts

Summarizing, the main contributions of this paper are the following:

- A novel individual-level network-based epidemic model to assess the impact of contact tracing.
- A rigorous estimation of the reproductive ratio *R*_0_ and confirmed case rate (infected to laboratory-confirmed transition) from observed confirmed case data.
- A thorough investigation of costs and benefits of contact-tracing in the reopening process in a college town of Kansas.

The individual-based network model is developed to represent the heterogeneity in people mixing. Our individual-based network epidemic model is general and flexible. It can be used to estimate, and model contact-tracing for COVID-19 in any location. It can also be used for any other disease that has a similar spreading mechanism like COVID-19.

This paper is organized as follows: section II proposes an individual-based contact network framework with two networks: the full network and the limited network, to represents the contact situation namely before the reopening process (under ‘stay-at-home’ order) and after the reopening process. Section III presents an epidemic model for the stochastic simulations of COVID-19 spreading. Section IV provides the implementation of the contact-tracing on a two-layer individual-based network framework and investigate the effectiveness of contact tracing in the reopening process. Finally, we provide a concluding remark of our research in section V.

## II. INDIVIDUAL-BASED CONTACT NETWORK MODEL

This section proposes a method to develop an individual-based contact network model capable of representing heterogeneous social mixing. In this network, occupied households are in the individual node level, a connection between two households represents the contact probability between members of these households. The network has *N* nodes and *n* people. To develop this network, we consider five age-ranges: under 18, 18 −24, 25 −34, 35 −59, and over 60. Each age-range has *n*_*i*_ people, where *i* ∈ {1, 2, 3, 4, 5}. We distribute the *n* people randomly into the *N* occupied households according to five social characteristics: age, average household sizes, family households, couple, living-alone^3^. We maintain the average household sizes, number of family households, number of couples, and number of living-alone households. Besides, a person under 18 years old is always assigned in a house with at least one adult person.

After assigning the people, an age-specific network is developed for each age range and a random mixing network for all ages. Then a combination of the six networks provides the full network. A full network represents a contact network for a typical situation. The configuration network model^8^ is used to develop age-specific networks and the random mixing network. The steps to develop age-specific networks are:

Step 1: For each person *j* (here, *j* ∈ 1, 2, …, *n*), contacts *c*_*j*_ is assigned from a Gaussian distribution 𝒩 (*µ, σ* ^2^). The mean *µ* of the Gaussian distributions are taken from the daily average number of contacts per person in each age-range^9–11^. The average daily contacts per person are given in Table I. For an under 18-year-old person, the number of contacts is assigned randomly from the 𝒩 (13.91, 6.95) distribution. For a person in 18 − 24 years age, the number of contacts is assigned randomly from the 𝒩 (21.25, 10.62) distribution. For a person in 25 − 34 years age, the number of contacts is assigned randomly from the 𝒩 (21.3, 10.65) distribution. For a person in 35 − 59 years age, the number of contacts is assigned from the 𝒩 (20.912, 10.46) distribution. For an over 60-year-old person, the number of contacts is assigned randomly from the 𝒩 (10.7, 5.35) distribution. In the random-mixing-network, the number of contacts is assigned randomly from the 𝒩 (2, 1) distribution for a person *j*. The Gaussian or normal distribution is the distribution of real numbers; therefore, the number from the 𝒩 (*µ, σ* ^2^) distribution is rounded to the closest integer.

**TABLE I.** Properties of the Age-specific-networks of the Manhattan, KS.

| Age-range | under 18 | 18-24 | 25-34 | 35-59 | over 60 |
| --- | --- | --- | --- | --- | --- |
| population | 8074 | 20378 | 9887 | 10581 | 6567 |
| average daily contacts per person<sup>9</sup> | 13.91 | 21.25 | 21.3 | 20.912 | 10.7 |
| average daily contacts with non-household members per person | 12.00 | 20.001 | 19.98 | 19.00 | 7.05 |
| % of neighbors in the same age-specific networks<sup>10</sup> | 85.63 | 90.48 | 90.29 | 84.95 | 71.43 |
| Number of edges in the age-specific networks | 40466 | 187723 | 88806 | 90835 | 16511 |

Step 2: For each person *j*, contacts for its belonging household *k* is assigned by (*c*_*j*_ − *h*_*k*_ − 1). Here, *c*_*j*_ is the number of contacts for a person *j, h*_*k*_ is the household size or number of people of the household *k*, person *j* lives in the household *k, j* = 1, 2, 3……*n*, and *k* = 1, 2, 3 *N*.

Step 3: From the mixing patterns of different age-ranges, people have a strong tendency to meet people with their same age range (more than 80%)^9–11^. Therefore, We keep the maximum number of contacts among the same age ranges and a small percentage for the other age ranges. The percentage of contacts in the same age-specific-network for each age-range is given in Table I. Degree *d*_*ki*_ of a node *k* in the age-specific network *i* is *s*% of (*c*_*j*_ − *h*_*k*_ − 1), here, *s*% of average daily contacts of a person happens with the people of his same age-range.

Step 4: After assigning degree, *d*_*ki*_ for *N* nodes or households, The configuration network model^8^ creates half-edges for each node, then chooses two nodes randomly and connect their half-edges to form a full edge^8^.

The population and network characteristics for the five age-specific networks for Manhattan, KS are given in Table I. According to census 2018, Manhattan, KS has *n* = 55, 489 people and *N* = 20, 439 occupied households^3^.

The full network is a combination of five age-specific networks and a random-mixing network. Adjacency matrix for the full network *A*_*f*_ is a summation of six adjacency matrices: 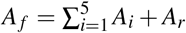. Here, *A*_*i*_ is the adjacency matrix for the age-specific network *i*, and *A*_*r*_ is the adjacency matrix for the random mixing network. Age-specific networks and the random mixing network are unweighted and undirected. However, the full network is a weighted and undirected network. The full network for Manhattan (KS) has 445, 350 edges. The average node degree for an individual household in the full-network is 43.647, and for an individual person is 16.0518 (which is consistent with^9^). The degree distribution is presented in Fig. 1. The networks are given in the supplementary materials.

**FIG. 1.**
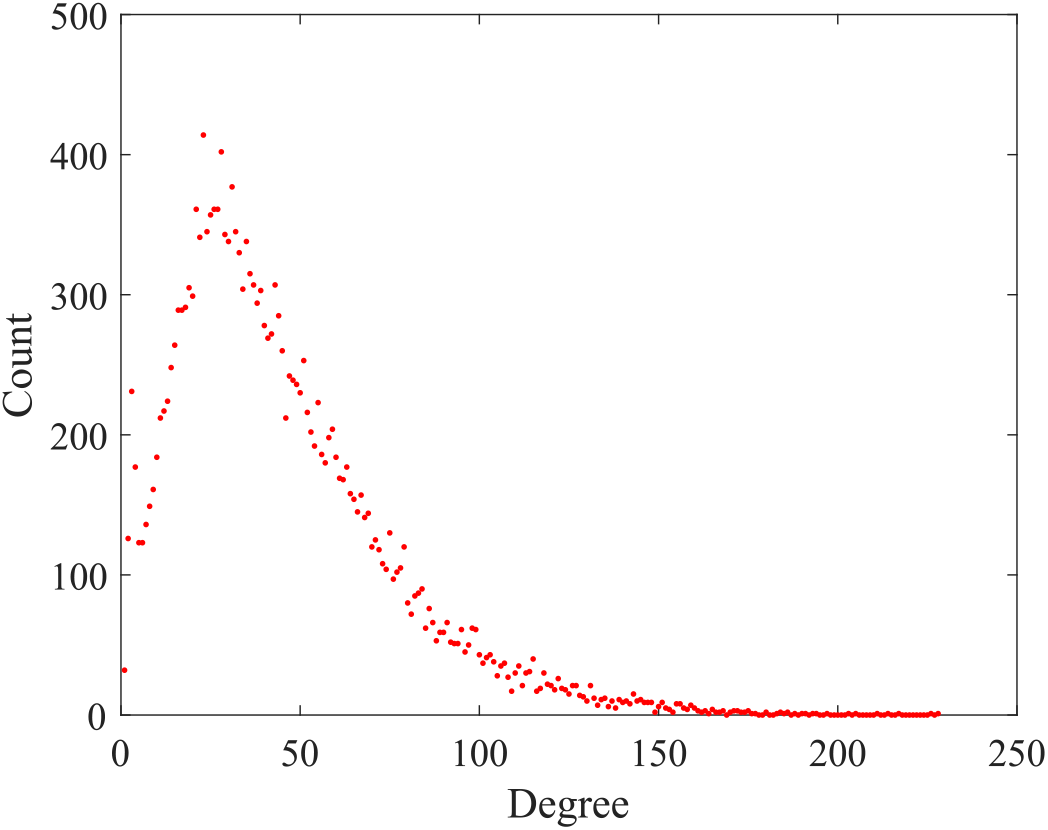
Degree distribution of the full network. In the network, households are at the node level. The network has 20, 439 nodes and 445, 350 edges. The average degree of this network is 43.647. The maximum degree in the network is 227.

The full network is a contact network in the normal situation; we modify it to represent the contact network in the pandemic lockdown; we name it limited network. Manhattan, KS, is the home of Kansas State University. Most of the people living in Manhattan, KS are closely related to Kansas State University, which is closed since early March 2020. Besides, Manhattan, KS was under the “Stay-At-Home” order from March 27, 2020 to May 4, 2020^12^. To represent this unusual situation, the full network is modified to a limited network version. As the educational institute was closed, we randomly reduce 90% links from the age-specific networks for the age-ranges under 18, and 18 − 24. The Google COVID-19 community mobility reports provide a percentage of movement changes in different places (for example, workplaces, recreational areas, parks)^13^. We reduced 40% links randomly from the age-specific networks for 25 − 34, and 35 − 59 age-ranges for the movement changes in the workplaces^13^. The number of links in the limited network is 155762. The limited network is given in the supplementary materials.

## III. EPIDEMIC MODEL

In this section, we design an epidemic model to simulate the COVID-19 spreading; later, we estimate the unknown parameters (reproductive ratio *R*_0_ and confirmed rate *δ*_2_) of the epidemic model. We simulate four reopening scenarios using the estimated parameters: under “Stay-At-Home” order, 25% reopening, 50% reopening, and 75% reopening. This model assumes that there is no particular mitigation strategies have applied except general lockdown.

### A. Susceptible-exposed-infected-confirmed (SEIC) epidemic model

This research propose a susceptible-exposed-infected-confirmed (SEIC) epidemic model to simulate the spreading of COVID-19 (Fig. 2). This model has four compartments: susceptible *S*, exposed *E*, infected *I*, confirmed *C*. A susceptible node does not introduce to the virus yet, an exposed node introduces to the virus, but the viremia level is not strong enough to infect others, an infected node has strong viremia to infect others, and a confirmed node is a laboratory-confirmed COVID-19 case. The SEIC model has three transitions, which are divided into two categories: edge-based (*S* → *E*), and nodal (*E* → *I*; *I* → *C*) transitions^14,15^.

**FIG. 2.**
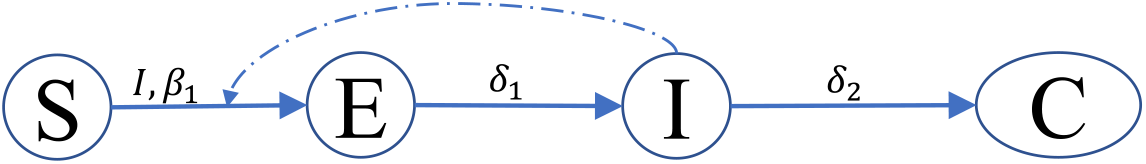
Node transition diagram of the susceptible-exposed-infected-confirmed (SEIC) epidemic model. This model has four compartments: susceptible (*S*), exposed (*E*), infected (*I*), and confirmed (*C*) compartments. The SEIC model has three transitions (presented by solid lines): *S* → *E* (edge-based), *E* → *I* (nodal), and *I* → *C* (nodal). The infected (I) compartment is the influencer compartment of the edge-based *S* → *E* transition. The dashed line presents the influence of the *I* compartment on the *S* → *E* transition. We estimate *R*_0_ and *δ*_2_ transition rate from data. we deduce *β*_1_ from *R*_0_.

An edge-based transition of a node depends on the state of its contacting nodes or neighbors in the contact network with its own state. A nodal transition of a node only depends on the own state. Each edge-based transition has an influencer compartment. A transition from susceptible to exposed (*S* → *E*) of a susceptible node depends on the infected neighbors of that node. Therefore it is an edge-based transition, and the infected compartment is the influencer compartment of this transition. In this work, we are using the term ‘neighbors of a node *k*’ for the nodes, which have the shortest path length 1 from the node *k*. The transition rate of the susceptible to exposed (*S* → *E*) transition of a node *k* is 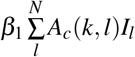, here, *A*_*c*_ is the adjacency matrix of the contact network, if *l* node is infected then *I*_*l*_ = 1 otherwise *I*_*l*_ = 0, and 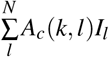 is the number of infected neighbors of the node *k*. The transition rate for the transition exposed to infected (E → *I*) is *δ*_1_. The confirmed rate of an infected person is *δ*_2_. We assume that a laboratory-confirmed case will be isolated and cannot spread the disease outside of his household anymore. An infected node will spread the disease for an exponentially distributed period with an expected value of 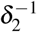. The unknown infected cases are present indirectly in our model through infected to confirmed transition. An unknown infected case will not be detected as a COVID-19 patient. If the estimated time for the infected to confirmed transition is higher than the average, it indicates that the system has more COVID-19 patient than laboratory-confirmed cases. A detail of the SEIC epidemic model is stated in Table II.

**TABLE II.**
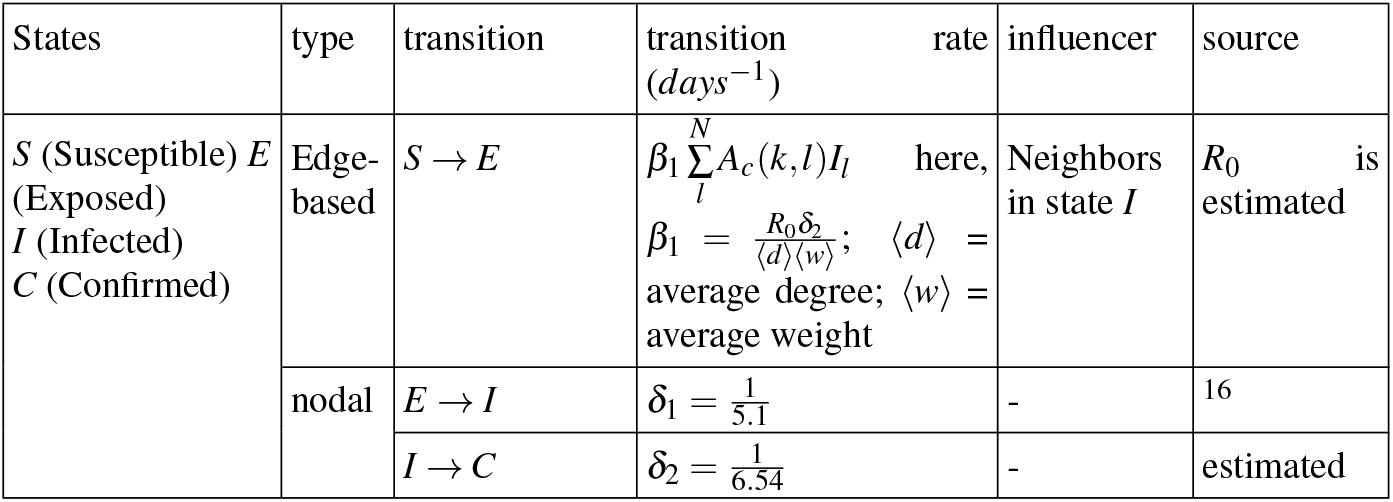
Description of the susceptible-exposed-infected-confirmed (SEIC) epidemic model.

### B. Stochastic simulation

To do the simulation, we use GEMFsim; it is a stochastic simulator for the generalized epidemic modeling framework (GEMF), which was developed by the Network Science and Engineering (NetSE) group at Kansas State University^17^. The GEMFsim is a continuous-time, individual-based, numerical simulator for the GEMF-based processes^14^. The network and epidemic model is the input of the GEMFsim, and the time dynamic of each node state is the output. In GEMF, the joint state of all nodes follows a Markov process that arises from node-level transition. A node can change its state by moving from one compartment to another compartment through a transition. One assumption of the GEMF system is, all the events or transitions are independent Poisson processes with the constant rate; this assumption leads the system to a continuous-time Markov process. Initially, the simulation starts by setting two infected nodes randomly.

### C. Parameter estimation for the SEIC epidemic model

The SEIC model has two unknown parameters: reproductive ratio *R*_0_, and confirmed or reporting rate *δ*_2_. To estimate the *R*_0_ and *δ*_2_, we have used confirmed cases in Riley County (Kansas) from March 25, 2020 to May 4, 2020. In this period, Kansas State University was closed, and ‘Stay-At-Home’ order was there. For the simulation of this period, a limited network is used (explained in section II), which is a modified version of the Full network to simulate the particular situation under the “Stay-At-Home” order.

The estimated value for *R*_0_ is 0.71 (95% confidence interval: 0.702 − 0.724) and for reporting rate *δ*_2_ is 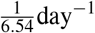 (95% confidence interval: 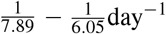). These estimated values are specific for Manhattan, KS. We have considered that some people will develop severe symptoms, and they will be reported as a confirmed case of COVID-19 sooner. However, some people will produce deficient symptoms, and may they will not be tested. Therefore, the estimated confirmed rate is an average of all possibilities. We use approximate Bayesian computation based on sequential Monte Carlo sampling (ABS-SMC) approach to estimate the parameters^18,19^.

A sensitivity analysis for *R*_0_ and *δ*_2_ on the mean-squared error between confirmed cases data and simulated results is presented in Fig. 6. From the sensitivity analysis, the mean-squared error is low when the reporting time is high. It indicates undetected COVID-19 patients in the system. It means that an infected node needs to be infected for a longer time for the better fitting with the data. It also indicates that the testing of COVID-19 is not sufficient.

**FIG. 3.**
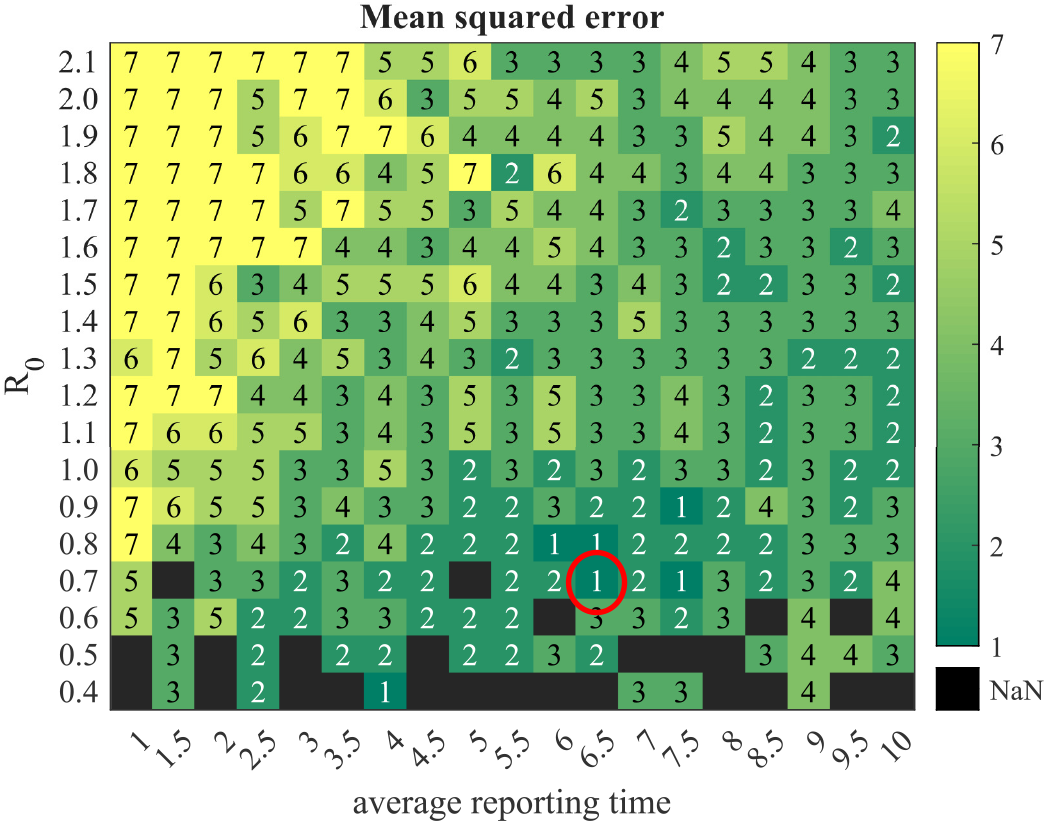
A sensitivity analysis. Mean-squared error (mse) between the time series of the confirmed cases (or cumulative new cases per day) of March 25, 2020 to May 4, 2020 and simulated results for a different combination of reproductive ratio and average reporting time (in days). The light-colored boxes represent more mse than dark-colored boxes. The color boxes with number “1” means that mse≤ 3, number “2” means that 3 <mse ≤ 10, number “3” means that 10 <mse ≤ 50, number “4” means that 50 <mse ≤ 100,number “5” means that 100 <mse ≤ 500,number “6” means that 500 <mse ≤ 1000,number “7” means that 1000 <mse. More than 80% times epidemic dies out in the combinations of the black squares, and confirmed cases are less than 10. The minimum error combination is showing by the red circle. We estimate *R*_0_ = 0.71 and average reporting time= 6.5 days.

**FIG. 4.**
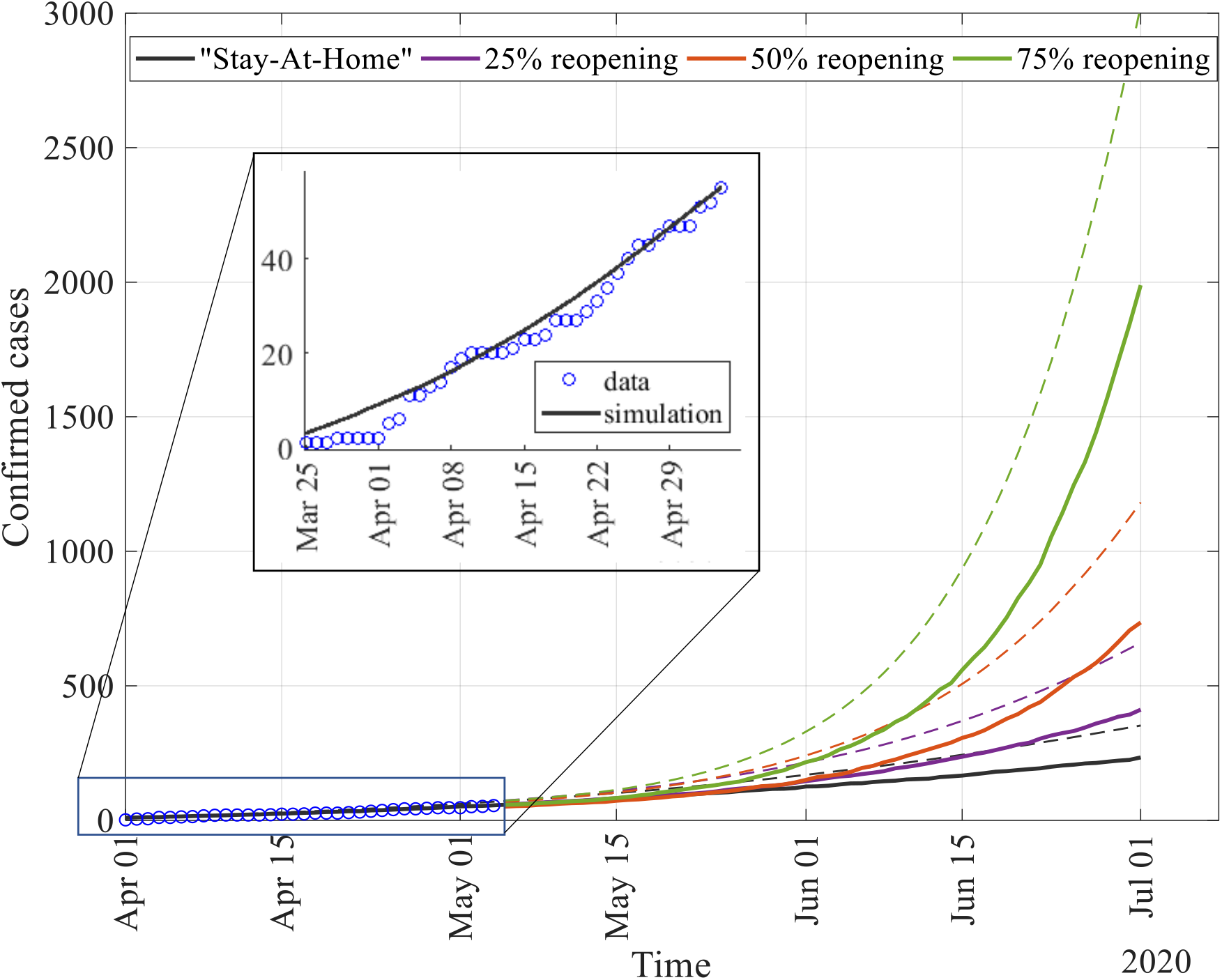
Confirmed cases in the four reopening scenarios after ‘stay at home’ order lifted on May 4, 2020. Solid lines represent the median, and dashed lines represent the mean of the 1000 stochastic realizations. The blue circle in the zoom-in window presents the confirmed case data of the COVID-19 in Manhattan (Kansas) for the time period from March 25, 2020 to May 4, 2020. We have used this time period to estimate the reproductive ratio and the average confirmed time.

**FIG. 5.**
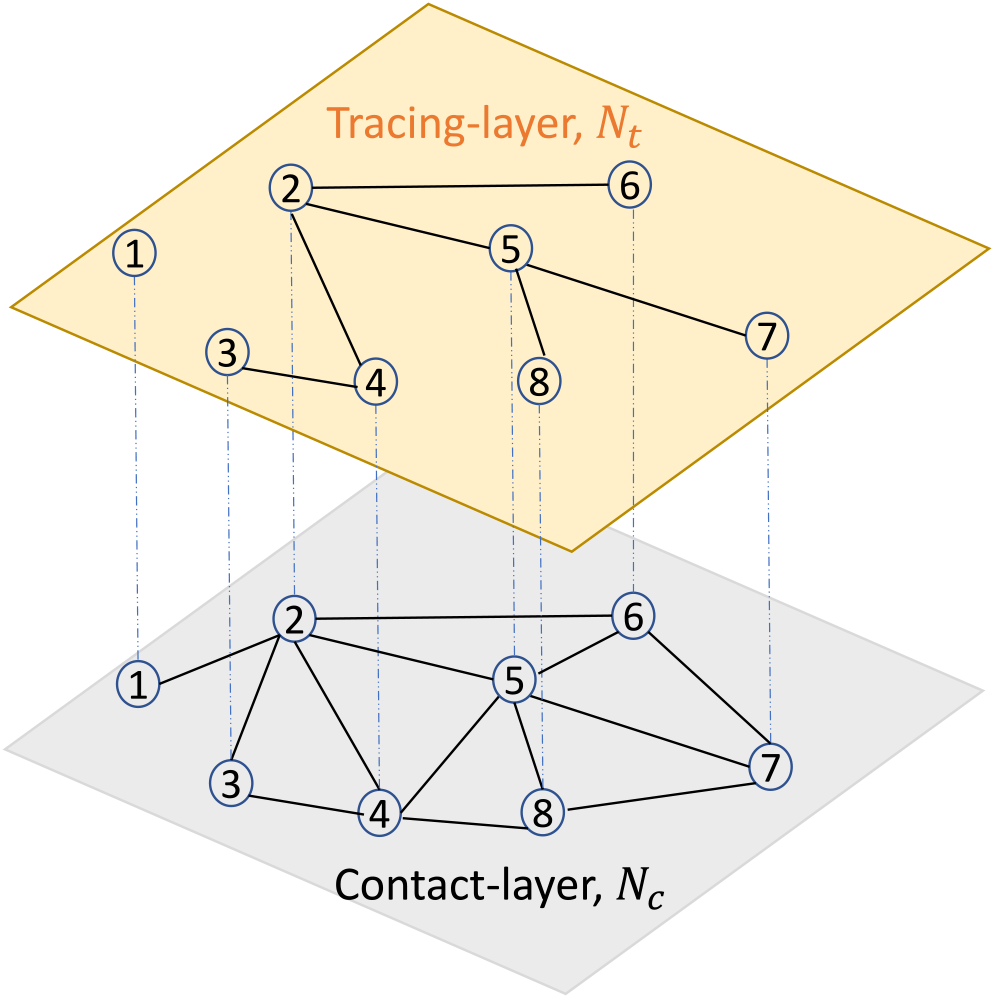
Two-layer network model: contact-layer *N*_*C*_, and tracing-layer *N*_*t*_. In this example, 50% of contacts of each node is traced; for example; node 4 has four neighbors in the contact network (2,3,5,8) however two neighbors in the tracing layer (2,3).

**FIG. 6.**
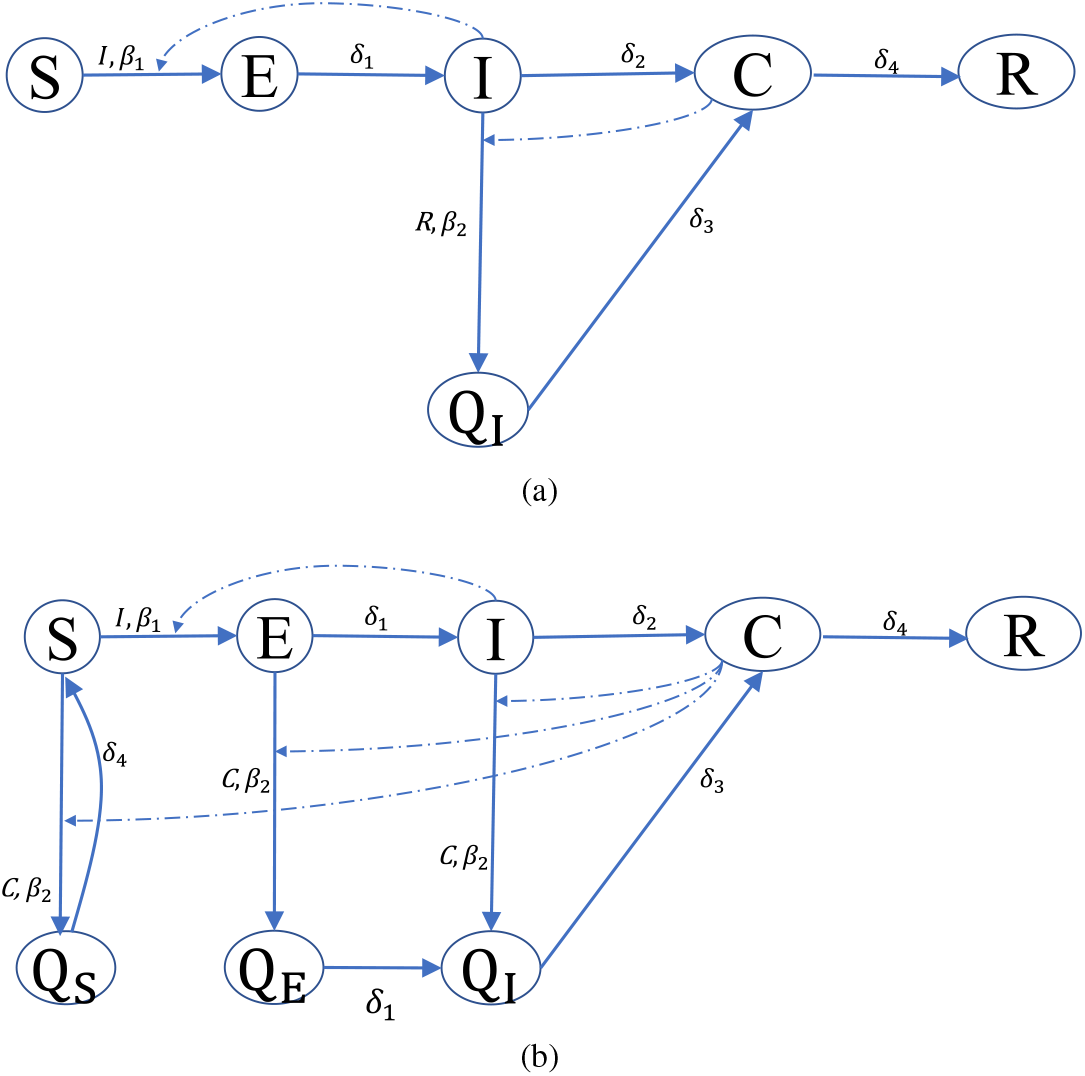
Node transition diagrams. a) SEICQ1 epidemic model, b) SEICQ2 epidemic model. The solid lines represent the node-level transitions, and the dashed lines represent the influence of the influencer compartment on an edge-based transition.

### D. Simulation in the reopening process without contact tracing

In this subsection, we simulate the confirmed cases (or cumulative new cases per day) for two months: June and July using the SEIC epidemic model with the estimated parameters. To simulate, we assume that there is no change except reopening from pandemic lockdown. We are presenting four reopening situations: Stay-at-home is still there or no reopening, 25% reopening, 50% reopening, and 75% reopening. Kansas has started to reopen step by step after May 4, 2020. We use the limited network to simulate from March 25, 2020 to May 4, 2020; then, we change the network concerning the reopening situation. For example, for a 25% reopening situation, we add 25% missing links randomly (which are present in the full network but not in the limited network). We preserve the states of each node at May 4, 2020 in the network then use it as the initial condition for the simulation for the reopening situation (from May 4, 2020 to July 1, 2020). Fig. 4 is showing the mean (dashed lines) and median (solid lines) of the confirmed cases of the 1000 stochastic realizations of the four reopening situations. The zoom-in window in Fig. 4 shows the time period when data was used to estimate the parameters of the epidemic model.

## IV. CONTACT TRACING

Contact tracing is a crucial mitigation strategy to control the spreading of COVID-19. In this section, we will implement contact tracing and observe the efficiency of the contact tracing in the different reopening scenarios. To implement contact tracing, we modify the basic SEIC epidemic model and propose a two-layer network model. In the implementation of the contact tracing, we follow the CDC’s guidance for contact tracing^6^.

### A. Two-layer individual-based network model

This work implements contact tracing in a two-layer network model: the contact network is in the first layer, and the tracing network is in the second layer (Fig. 5). We will call the first layer as the contact-layer and second layer as the tracing-layer in the rest of the paper. In the *t*%-tracing-layer, *t*% of links of each node in the contact-layer are preserved randomly to form tracing-layer (A 50% tracing-layer is presented in Fig. 5). The contacts/neighbors of a confirmed (C) node in the tracing-layer will be tested and quarantined.

### B. Epidemic model for contact tracing

For the contact tracing mitigation strategy, we consider two approaches for isolation: I) only infected neighbors of a confirmed case in the tracing layer will be isolated, II) all the neighbors of a confirmed case in the tracing layer will be isolated. For the case I, we propose the SEICQ1 epidemic model, and for case II, we propose the SEICQ2 epidemic model.

The SEICQ1 model has six compartments: susceptible (*S*), exposed (*E*), infected (*I*), confirmed (*C*), quarantined-infected (*Q*_*I*_), and removed (*R*). The SEICQ2 model has eight compartments: susceptible (*S*), exposed (*E*), infected (*I*), confirmed (*C*), quarantined-susceptible (*Q*_*S*_), quarantined-exposed (*Q*_*E*_), quarantined-infected (*Q*_*I*_), and removed (*R*). The transitions *S* → *E, E* → *I*, and *I* → *C* are the same as the base SEIC model.

In the SEICQ1 model, neighbors of a confirmed node in the tracing-layer will be monitored, and infected neighbors will go to the quarantined-infected (*Q*_*I*_) state immediately with rate *β*_2_, therefore infected to quarantined-infected (*I* → *Q*_*I*_) transition is an edge-based transition and confirmed compartment is the influencer of this transition. Here, *β*_2_ ≥ 1. A COVID-19 positive neighbor of a confirmed node will go to the confirmed state immediately with *δ*_3_ rate, *Q*_*I*_ → *C* is a nodal transition. A confirmed node will be removed from the system with *δ*_4_ rate (here, 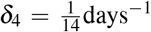), and its neighbors are not going to be monitored anymore after 14 days. A confirmed or removed node can not spread the disease anymore. The purpose of the transition *C* → *R* is to monitor the neighbors of a confirmed node for 14 days. The node transition diagram of the SEICQ1 model is given in Fig. 6a. A description of the 6 transitions of the SEICQ1 model is given in the Table III.

**TABLE III.**
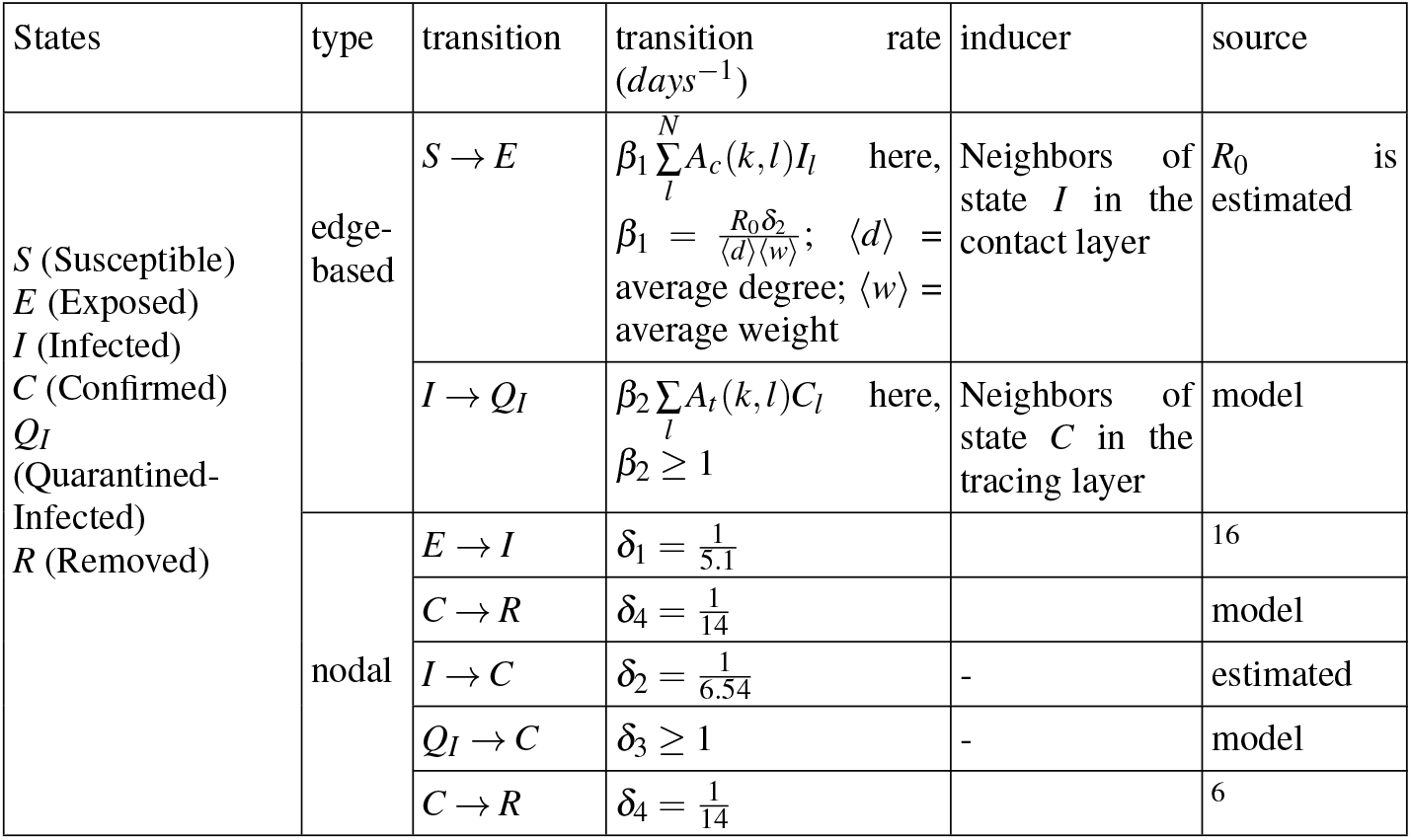
Description of the SEICQ1 epidemic model.

In the SEICQ2 model, neighbors (susceptible, exposed, and infected) of a confirmed node in the tracing-layer will be monitored and quarantined. The SEICQ2 model has four new transitions than the SEICQ1 model: susceptible to quarantined-susceptible (*S* → *Q*_*S*_), exposed to quarantined-exposed (*E* → *Q*_*E*_), quarantined-exposed to quarantined-infected (*Q*_*E*_ → *Q*_*I*_), and quarantined-susceptible to susceptible (*Q*_*S*_ → *S*). The SEICQ2 model is presented in Fig 6b. A description of the 10 transitions of the SEICQ2 model is given in Table IV.

**TABLE IV.**
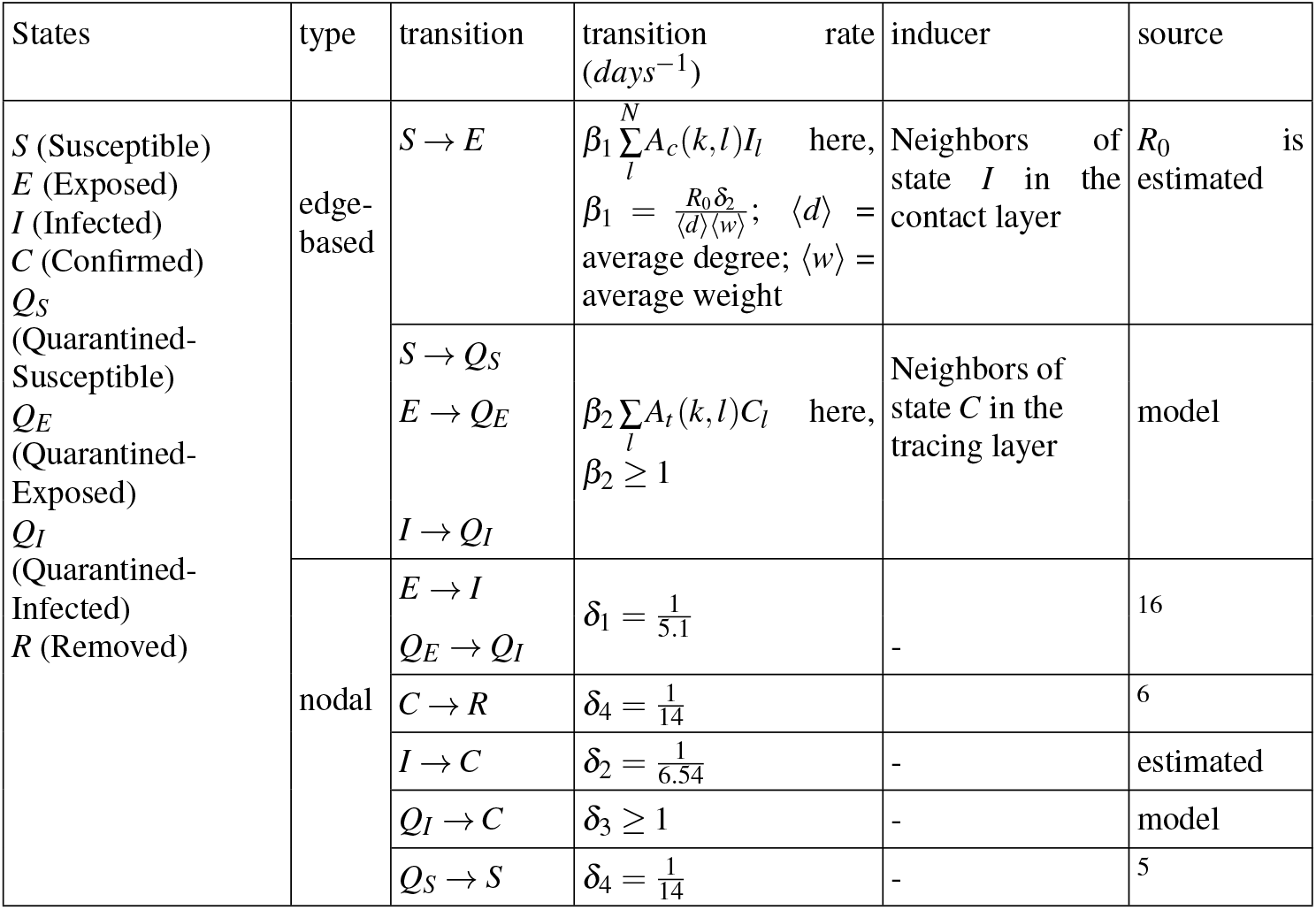
Description of the SEICQ2 epidemic model.

### C. Impact of contact tracing

Contact tracing can minimize the effect of the reopening process and control the spreading of COVID-19. We apply contact tracing after May 4, 2020 in Manhattan, KS. The plot of confirmed cases on Jul 1, 2020 is presented in Fig. 7 for four reopening situations : 25% reopening, 50% reopening, 75% reopening, and 100% reopening for the different levels of contact tracing. The dashed lines in Fig. 7 represents the mean, and solid lines represent the median of the 1000 stochastic realization for the SEICQ1 and SEICQ2 model.

**FIG. 7.**
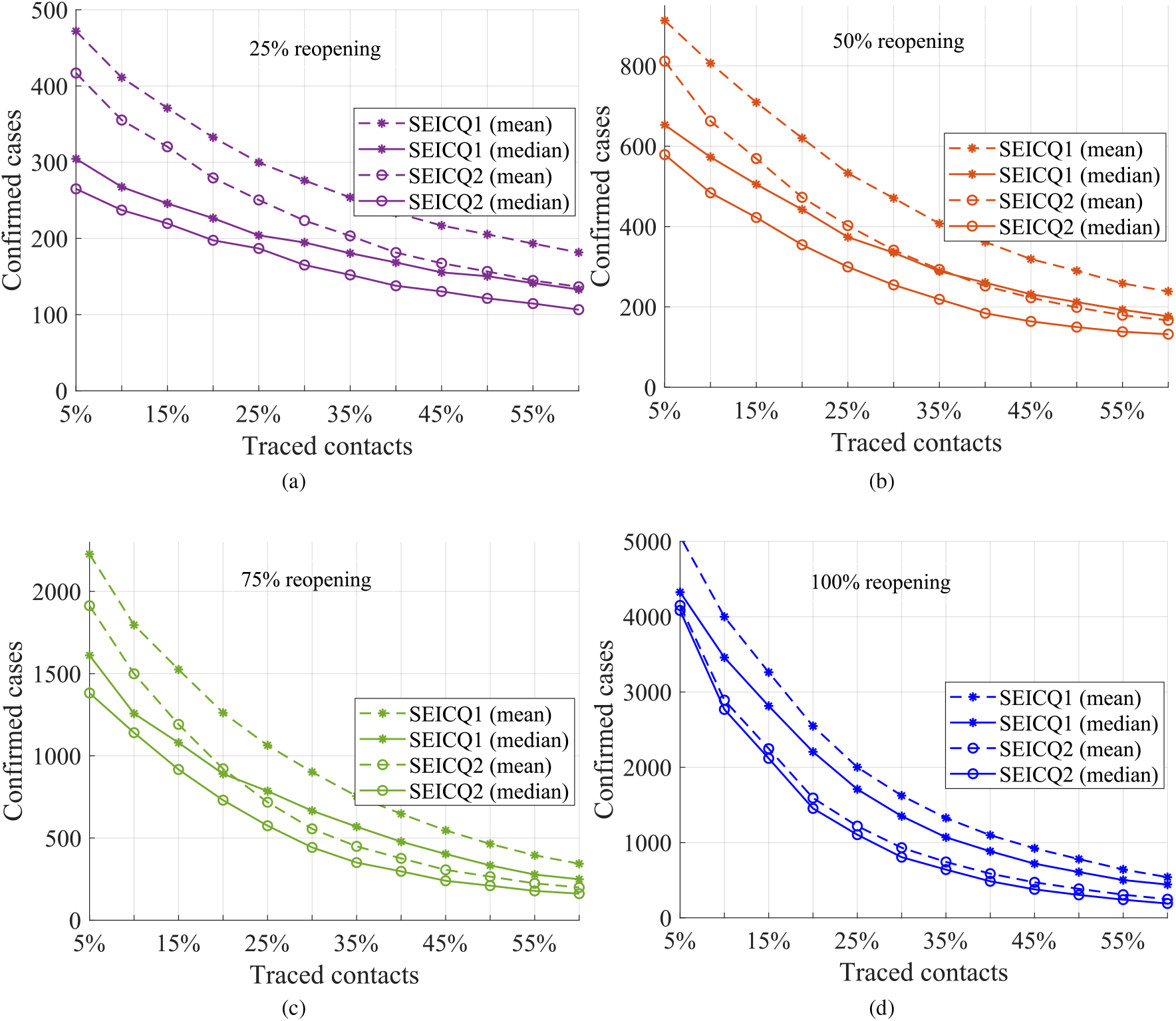
Impact of contact tracing. Total reported cases in two months after ‘Stay home order’ lifted for different movement restrictions scenarios. Contact tracing is applied after May 4, 2020. This figure is showing the median (solid lines) and mean (dashed lines) value of 1000 stochastic realizations.

The difference between SEICQ1 and SEICQ2 is that SEICQ1 quarantines only the infected neighbors of a confirmed case in the tracing layer however SEICQ2 quarantines susceptible, exposed, and infected neighbors of a confirmed case in the tracing layer. The SEICQ2 model is always efficient than the SEICQ1 epidemic model. However, both approaches can reduce the number of confirmed cases, even in the 100% reopening situation. For any reopening situations, tracing more than 60% of the contacts in the SEICQ2 can reduce the median of the 1000 stochastic realizations of the confirmed cases more than 96.5% on July 1, 2020 and in the SEICQ1 can reduce the median of the 1000 stochastic realizations of the confirmed cases more than 92%. The SEICQ2 model can reduce the confirmed cases on July 1, 2020 for more than 75% in the 25% reopening, more than 82% in the 50% reopening, more than 92% in the 75%, and more than 96% in the 100% reopening with compare to no-contact-tracing (SEIC model).

The SEICQ2 model can reduce the reported cases further compared to SEICQ1 for the same amount of contact tracing (Fig. 7). However, the SEICQ2 model has a drawback; it isolates susceptible persons. The number of total quarantined susceptible households in the simulation time period for different amounts of traced contacts for the SEICQ2 model is presented in Fig. 8. The quarantined susceptible households increase with the increase of tracing; however, after tracing 20% of contacts, the quarantined susceptible households start to decrease with the increase of tracing (Fig. 8). If we consider quarantined susceptible households are the cost of SEICQ2 model, then it is cost-effective to trace contacts of the confirmed cases more than 20%. The possible reason for decreasing the number of quarantined households with the increasing of contact-tracing after 20% is the smaller number of the infected cases or the smaller epidemic size. Although each confirmed case will give a long list of possible contacts, this effect will be balanced out by a decreasing number of the confirmed cases.

**FIG. 8.**
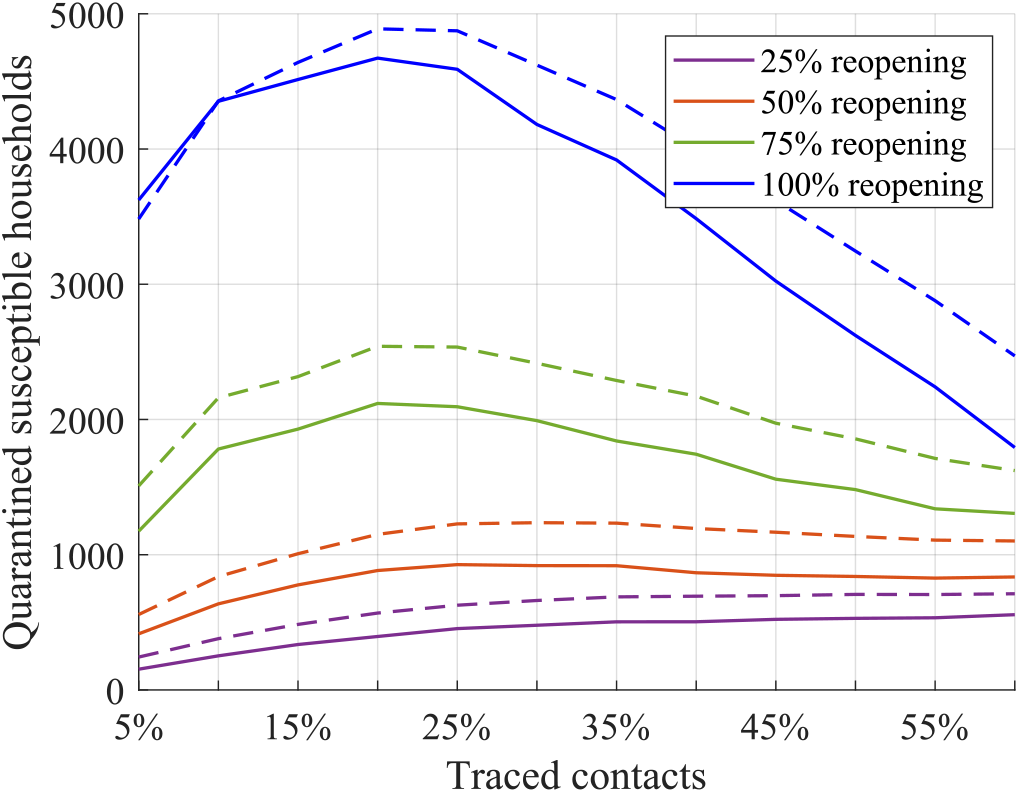
Total quarantined susceptible household in two months after May 4, 2020 for SEICQ2 epidemic model for the four reopening situations.This figure is showing the median (solid lines) and mean (dashed lines) value of 1000 stochastic realizations.

## V. CONCLUSION

This research studies contact tracing as a key mitigation strategy to control the COVID-19 transmission in the reopening process of a college town in the rural region of the USA. Therefore, we propose a general framework to develop an individual-based contact network epidemic model to estimate parameters and implement contact tracing. This model is used to estimate the reproductive ratio (*R*_0_) and confirmed rate (*δ*_2_) in Manhattan, KS, for the COVID-19 spreading.

The individual-based network model represents the heterogeneous mixing nature of a population. To investigate transmission at the individual level, we develop an individual-based contact network model where households are presented by network nodes. The contact network is a combination of five age-specific networks and one random-mixing network; this approach allows us to change an age-specific network according to any change in the society (for example, summer break, pandemic lockdown). The pandemic lockdown reduces the contacts mostly among the people who are students. Therefore, age-specific networks for under 18 and 18-24 changed. Pandemic lockdown also affects people in 25-34, 35-59 age-ranges. We propose a ‘full network’ to represent the usual situation; then, we modify the age-specific networks of the full network to represent pandemic lockdown. The modified network is the limited network, a reduced version of the full network. The average degree of the full network is 43.647 for Manhattan, KS which means that each household has probable direct connections with an average of 43.647 households. The full network is connected and provides an approximation of the contact network at the household level, which is useful for doing the simulation anonymously.

We propose a susceptible-exposed-infected-confirmed (SEIC) epidemic model in the limited network to simulate COVID-19 transmission from March 25, 2020 to May 4, 2020. We estimate the unknown parameters of the SEIC model for the Manhattan, KS, using approximate Bayesian computation based on sequential Monte Carlo sampling. We use confirmed cases as an observed data set. Designing an optimal epidemic model to simulate epidemic spreading is essential. However, it is challenging to design an epidemic model for COVID-19 spreading with limited knowledge; understanding the spreading of COVID-19 needs more investigation. Asymptomatic carriers of the SARS-CoV-2 are present in the spreading of COVID-19^20^. However, more research is needed to get information about how much an asymptomatic case can transmit the SARS-CoV-2 virus. Asymptomatic cases are included in our model indirectly. Concerning the unclear role of immunity, we assume that the immunity of a recovered COVID-19 patient is not going to fade in the short period analyzed in our simulations. In addition, it is important to keep the model simple, since the data available to estimate parameters is limited. Therefore, we propose a simple but dynamic and flexible epidemic model to simulate COVID-19 spreading, which has only two unknown parameters. The model can easily cope with additional information that may be available in the future.

The estimated reproductive ratio is much smaller in Manhattan, KS (estimated *R*_0_ = 0.71) because of the ‘Stay at home’ order. In Manhattan, 51% of people have age below 24 years, who get a chance to stay at home because of the online curriculum in educational institutions. However, the reproductive ratio will change when educational institutes start their in-person curriculum (in the 100% reopening *R*_0_ = 2.0301). There are 301 college towns in the USA^21^, which have a similar population structure like Manhattan, KS. A practical contact tracing approach can help to control the epidemic in those college towns.

We implement contact tracing by using a two-layer network model. We assess the impact of contact tracing in the four reopening situations: 25 % reopening, 50 % reopening, 75 % reopening, and 100 % reopening. Reopening without vaccination can produce more infected cases. It is essential to access the efficacy of the contact tracing in the reopening path. Our investigation indicates that more than 50% contact tracing can control the spreading of COVID-19 even in the 100% reopening situation. The number of quarantined susceptible people increases with the increase of traced contacts, however after 20%, the number of quarantined susceptible people decreases with the increases of the traced contacts. We consider that quarantined susceptible people represent the cost of contact tracing with a quarantined strategy. Therefore it is cost-effective to trace more than 20% contacts of a confirmed case. Our investigation indicates that a sufficient amount of contact tracing can reduce the impact of COVID-19 spreading in the reopening process of a location where the epidemic is in an initial stage. At first, the quarantined susceptible people increase with the percentage of traced contacts, however after a certain amount of traced contacts, the quarantined susceptible people start to decrease with the increase in the percentage of traced contacts.

## Data Availability

n/a

## ACKNOWLEDGMENT

This work has been supported by the National Science Foundation under Grant Award IIS-2027336.

## VI. DATA AVAILABILITY

The data that supports the findings of this study are available within the article [and its supplementary material].

